# Vitamin D Insufficiency and Respiratory Muscle Dysfunction in Myotonic Dystrophy Type 1 - A Prospective Cohort Study

**DOI:** 10.64898/2026.11.17.26368226

**Authors:** Daniel Jaraj, Ólöf Elíasdóttir, Stefan Winblad, Christopher Lindberg

**Author notes:** Correspondence: Daniel Jaraj, MD PhD, Sahlgrenska University Hospital, Neuromuscular unit, Institute of Neuroscience and Physiology, Gothenburg Sweden.

## Abstract

**Introduction:** Respiratory complications are a leading cause mortality in myotonic dystrophy type 1 (DM1), making it essential to identify modifiable factors that influence long-term pulmonary function. Vitamin D is known to have an important role in muscle physiology, but its potential relevance for respiratory muscle function has previously not been examined in this patient population.

**Methods:** This was a prospective cohort study including patients with genetically verified DM1 (n = 127) aged 18 years or more. Mean follow-up was 4.3 years. All patients underwent a series of examinations including blood tests and measurement of Vitamin D levels and Forced vital capacity % of predicted (FVC%).

**Results:** Vitamin D insufficiency (<50 nmol/L) was present in 43% of participants. Mean FVC% decreased modestly from 65.7% to 63.6% during follow-up (p=0.004). Patients with vitamin D insufficiency had significantly lower FVC% at follow-up compared to those with sufficient levels (54.7% vs 71.2%, p<0.001), and a higher proportion had worsened FVC% (72% vs 50%, p=0.04). In multivariable regression analyses adjusted for disease severity, lower vitamin D levels were independently associated with worse FVC% at follow-up (β=0.12, p=0.04). When vitamin D was examined as a categorical variable in an adjusted model, patients with insufficiency had on average 4.2 percentage points lower FVC% at follow-up compared to those with sufficient levels (β=−4.17, p=0.018), independent of baseline lung function and disease severity.

**Discussion:** Lower baseline Vitamin D levels were strongly related to lung function at follow-up in patients with DM1. These findings remained significant after adjusting for potential confounders. However, because FVC% remained largely stable during follow-up, the possibility to assess the long-term effects of vitamin D on change in respiratory function was limited.

## Introduction

Respiratory complications are one of the leading causes of mortality in Myotonic Dystrophy type 1 (DM1)^1^. Optimizing pulmonary function and preventing decline in ventilatory capacity is therefore essential for effective patient management^2^.

Muscle weakness, disease severity and longer disease duration are known risk factors for decline in lung function in DM1, but there is little data on risk factors that are potentially modifiable ^3, 4^. In current management of DM1, inspiratory muscle training, non-invasive ventilation, smoking cessation, and weight reduction in cases of overweight or obesity are well-established interventions^2^. However, to the best of our knowledge, no additional modifiable risk factors have yet been identified.

Vitamin D has an integral role in skeletal muscle function through several pathways such as calcium homeostasis, intracellular signaling, protein synthesis and mitochondrial activity^5^. For example, mechanistic studies have shown that Vitamin D promotes muscle regeneration, as evidenced by upregulation of vitamin D-receptor expression after muscle injury ^6^, whereas vitamin D deficiency increases oxidative stress and reduces mitochondrial function in knock-out animals ^7^. At the same time, multiple clinical studies have found associations between low vitamin D levels and muscle weakness, and data from several randomized trials have demonstrated that vitamin D substitution improves muscle strength ^8–10^. There is also data, albeit more sparse, on the effects of reduced pulmonary function in patients with vitamin D deficiency, but evidence in neuromuscular cohorts remains limited^11, 12^.

Although patients with DM1 frequently have Vitamin D insufficiency, the relationship between vitamin D status and respiratory decline in this patient group is unclear^13^. Because of the clinical importance of detecting early deterioration in respiratory capacity, and the biological plausibility linking vitamin D insufficiency to impaired muscle function we aimed to evaluate the relationship between baseline vitamin D status and subsequent changes in forced vital capacity (FVC) among patients with DM1.

## Material and methods

### Study design and participants

The present study is a substudy of a prospective cohort study designed to investigate the natural history and clinical manifestations of DM1. A comprehensive standardised multidisciplinary assessment was performed at baseline and follow-up, encompassing neurological, functional, cognitive and laboratory evaluations, to enable investigation of multiple research questions. The present analysis focuses specifically on the association between vitamin D insufficiency and respiratory function over time. Baseline examinations were made in in 2018. According to the initial study protocol, two follow-up assessments were planned after 2 years and after 4 years respectively. However, because of the interruption of the Covid-19 pandemic only one follow-up examination was possible. This was done 3-4 years after baseline (on average 4.3 years). All assessments were conducted at at the Sahlgrenska university hospital - a specialized neuromuscular centre - using standardized clinical protocols.

All participants had a verified pathological CTG repeat expansion in the *DMPK* gene and were aged ≥18 years at enrolment. CTG repeat length was retrieved from each patient’s medical record and reflects the results of routine clinical genetic testing performed on peripheral blood according to standard laboratory procedures at the department of clinical genetics at our university hospital. For patients where repeat length was reported as an interval, the midpoint value was used. For patients where an exact value or minimum threshold was reported (n=14), the reported value was used directly. Study participants included patients with childhood-, juvenile-, adult- and late adult-onset DM1. Because the study comprised extended visits and comprehensive multidisciplinary testing including self-reporting of symptoms and cognitive testing, participation was deemed to be difficult and too strenuous for those with congenital DM1. This patient groups was therefore not included in the study. 127 patients underwent baseline examinations of which 102 (80 %) completed follow-up assessments. Mean follow-up time was 4.3 years (SD 0.76).

### Clinical assessments

All participants underwent comprehensive clinical evaluations by a multidisciplinary team including neurologists, physiotherapists, and occupational therapists. Standardized examinations included testing of strength and functional neuromuscular evaluation, cognitive screening, anthropometric measurements, ECG and laboratory exams. Pulmonary function was assessed using standardized spirometry performed by trained physiotherapists according to established praxis. Forced vital capacity (FVC) was measured in litres and recorded as percent of predicted values (FVC%) based on age, sex, and height according to Swedish reference values described by Hedenström et al^14^. To enable longitudinal analysis, all clinical, spirometry and laboratory assessments at follow-up were identical to baseline. Six-minute walk test (6MWT) was assessed by trained physiotherapist, whereas MIRS^15^ was determined by physicians who were specialists in clinical neurology subspecialized in neuromuscular disorders.

### Vitamin D status

Serum 25-hydroxyvitamin D (Vitamin D) concentrations were obtained through venous blood samples and measured in accordance with standard clinical chemistry analysis at our university hospital. Analyses were made using chemiluminescent Microparticle Immunoassay (CMIA) and values reported in units of nmol/L. For categorical analyses vitamin D insufficiency was based on accredited laboratory reference values and defined, per local guidelines, as serum concentration < 50 nmol/L. Data on medications was not collected and we were therefore unable to account for possible vitamin D-supplementation. Additionally, plasma levels of phosphate and ionized calcium were measured using photometry and ion selective electrode analysis. The reason for including phosphate and ionized calcium in the analyses was to analyze possible interactions given that they are part of the same metabolic pathways as vitamin D and also have homeostatic roles in muscle contractility^16^.

### Statistical analysis

Continuous variables were inspected for distributional properties using density plots. Group comparisons for continuous variables were made using independent samples t-tests or the Mann–Whitney U test, as appropriate. Associations between categorical variables were examined using the chi-square test.

The crude association between serum vitamin D levels and FVC% at follow-up was first examined by comparing means and proportions in those with and without insufficient levels as well as using univariable linear regression. Using vitamin D as a continuous predictor multivariable linear regression models were conducted to examine the association between vitamin D levels and FVC% at follow-up adjusting for potential confounders. Additionally, a multivariable linear regression model was also run using vitamin D status as a categorical predictor comparing sufficiency (≥50nmol/L) vs insufficiency (<50nmol/L). Covariate selection for the adjusted models was guided foremost by clinical relevance and causal diagrams: Directed Acyclic Graphs (DAGs)^17^ (figure shown in supplements). Accordingly, the minimal sufficient adjustment set that was identified included age, body mass index (BMI) and disease severity.

Because direct measures of disease severity was not possible, 6MWT, MIRS and CTG-expansion size were used as clinical proxies for disease severity, in the adjusted analyses. To examine change in FVC% we also performed the same multivariable model including baseline FVC% as a covariate.

Age, as well as plasma levels of Phosphate and ionized Calcium were tested as interaction terms. Model assumptions were assessed by inspection of standard diagnostic plots. No major violations were seen.

Missing data were handled using complete-case analysis. All statistical tests were two-sided, and a p-value <0.05 was considered statistically significant. Statistical analyses were conducted using R (R version 4.5.2; R Foundation for Statistical Computing, Vienna, Austria)^18^.

### Ethical considerations

Informed consent was collected from all participants prior to entering the study. The study was conducted in accordance with the Declaration of Helsinki (2000) and was approved by The Swedish Ethical Review Authority (number 826-16).

## Results

### Baseline characteristics

The cohort included 127 participants at baseline and 80 % (n = 102) completed follow-up examinations. Regarding DM1 subtypes, 38 (30 %) had childhood or juvenile onset, 67 (54 %) had adult onset and 19 (15 %) had late onset DM1. Mean follow-up time was 4.3 years (SD 0.76). Of the baseline participants, 52 % were women (n = 66) and 48 % men (n = 61). Mean age was 48 years (SD 14.2, range 21-80 years) and mean CTG expansion was 442.5 (SD 353.4). Median vitamin D level was 53 nmol/L (IQR 34) and 54 patients (43 %) had levels corresponding to insufficiency (< 50 nmol/L). Baseline characteristics are shown in table 1.

**Table 1.**
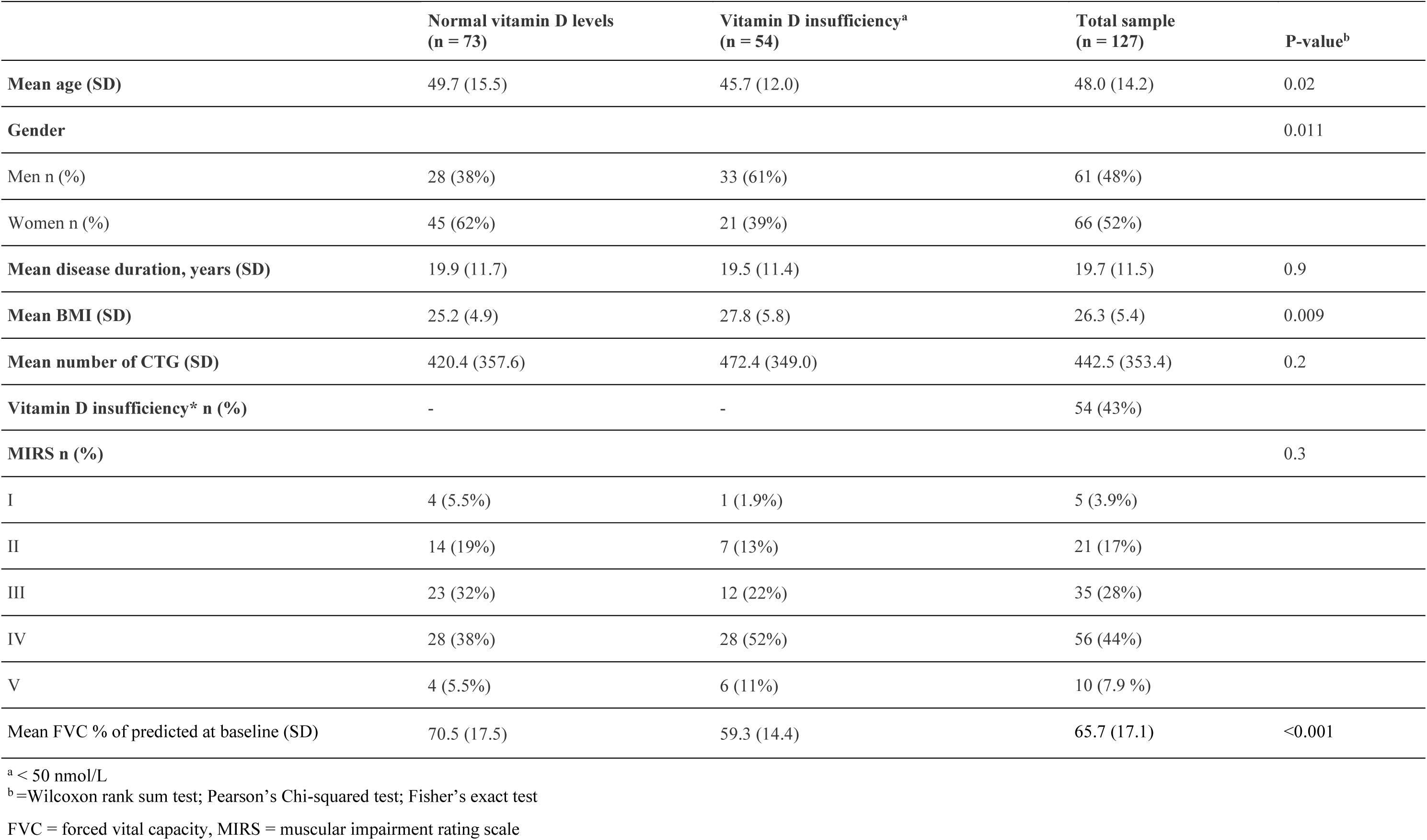
Baseline characteristics.

### Change in FVC and association with Vitamin D

Overall FVC% was relatively stable during the study period and decreased modestly: Mean baseline FVC% was 65.7 (SD 17) vs FVC% 63.6 (SD 19) at follow-up (p = 0.004, Cohens D = 0.12).

There was a strong univariate association between baseline Vitamin D levels and FVC% at follow up: Mean FVC% was lower among those with vitamin D insufficiency compared to those with sufficient levels (54.7 % vs 71.2 %, p<0.001), with a slight dose-response relationship when comparing tertiles of vitamin D levels (figure 1). Among those with baseline vitamin D insufficiency, a significantly higher proportion had worsened FVC% compared to those with normal vitamin D levels (72 % vs 50 %, p=0.04), shown in figure 2. In the univariate regression analyses, higher serum vitamin D levels were associated with higher FVC% at follow-up: β = 0.21 (p = 0.004). In a univariate model including only baseline FVC% as the sole predictor of follow-up FVC%, the beta coefficient for baseline FVC% was 1.003 (p < 0.001) indicating that lung function at follow-up was largely determined by baseline lung function.

**Figure 1.**
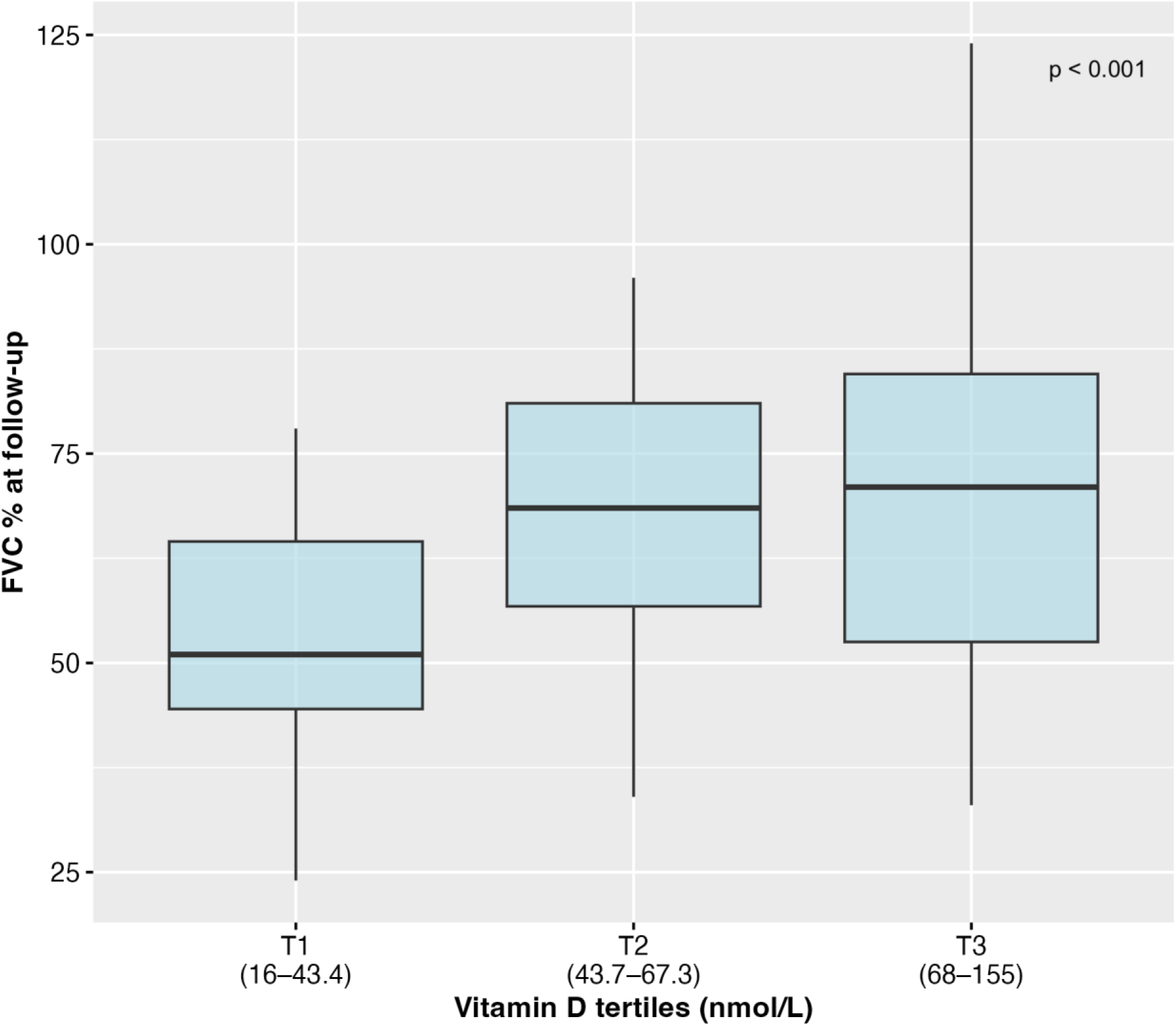
Median FVC% at follow-up by vitamin D tertiles

**Figure 2.**
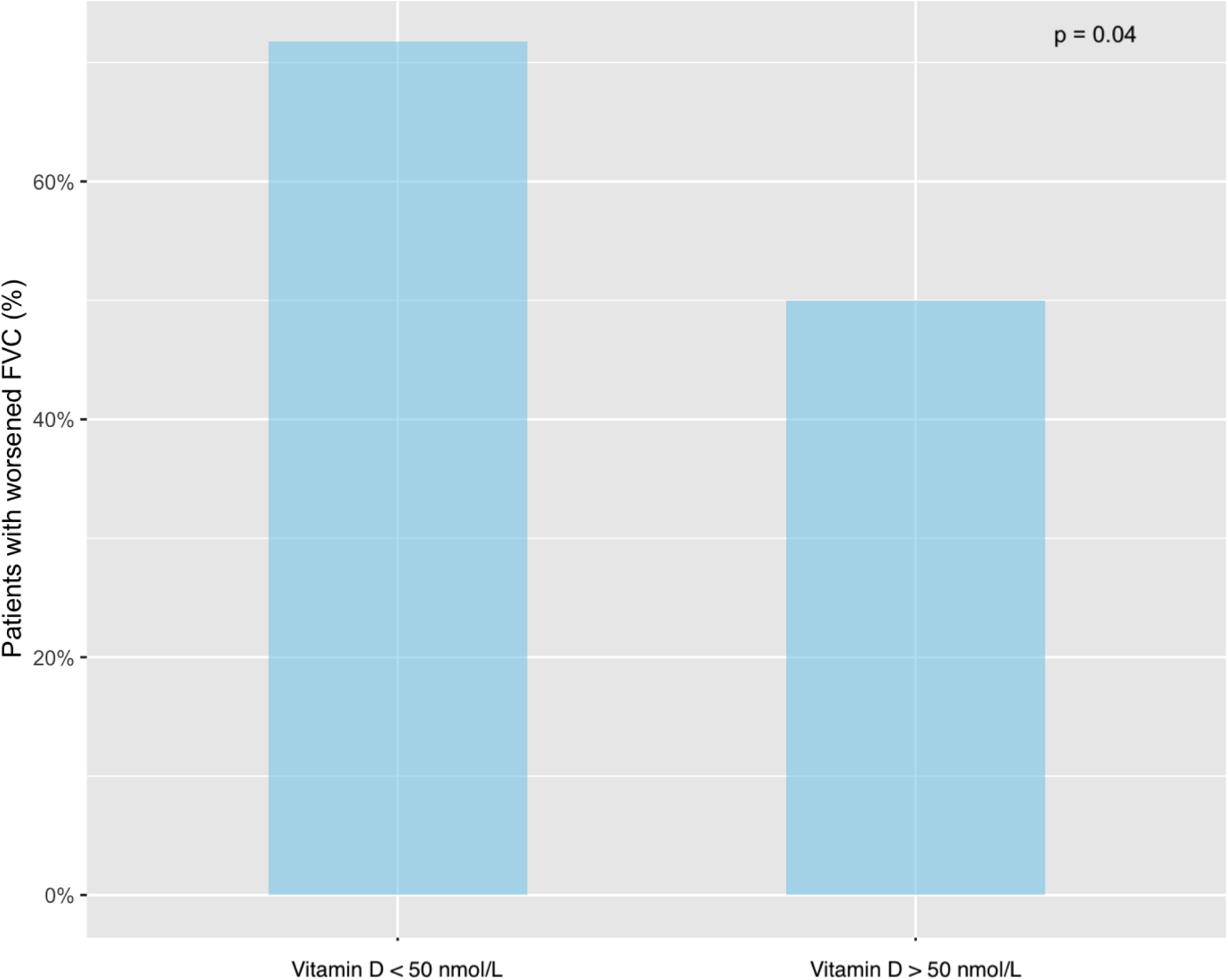
Proportion of patients with worse FVC% at follow-up comparing those with Vitamin D insufficiency vs those with sufficient levels

### Multivariable models with Vitamin D as continuous predictor

In the multivariable linear regression analysis, adjusting for age, BMI, baseline MIRS, 6MWT and CTG-repetitions, vitamin D was independently associated with FVC% at follow-up: β = 0.12 (p = 0.04). Baseline MIRS, 6MWT and CTG expansion size were also significantly associated with worse FVC%. In the model were baseline FVC% was included as a covariate (i.e modeling change in FVC%), the relationship with vitamin D did not remain statistically significant. Instead, baseline FVC% was the strongest predictor of follow-up FVC%: β = 0.81 (p < 0.001), adjusted R2 = 0.85, again indicating that most of the variation in follow-up FVC% was explained by baseline lung function. These results are shown in table 2. None of the tested interaction terms were found to be significant.

**Table 2.**
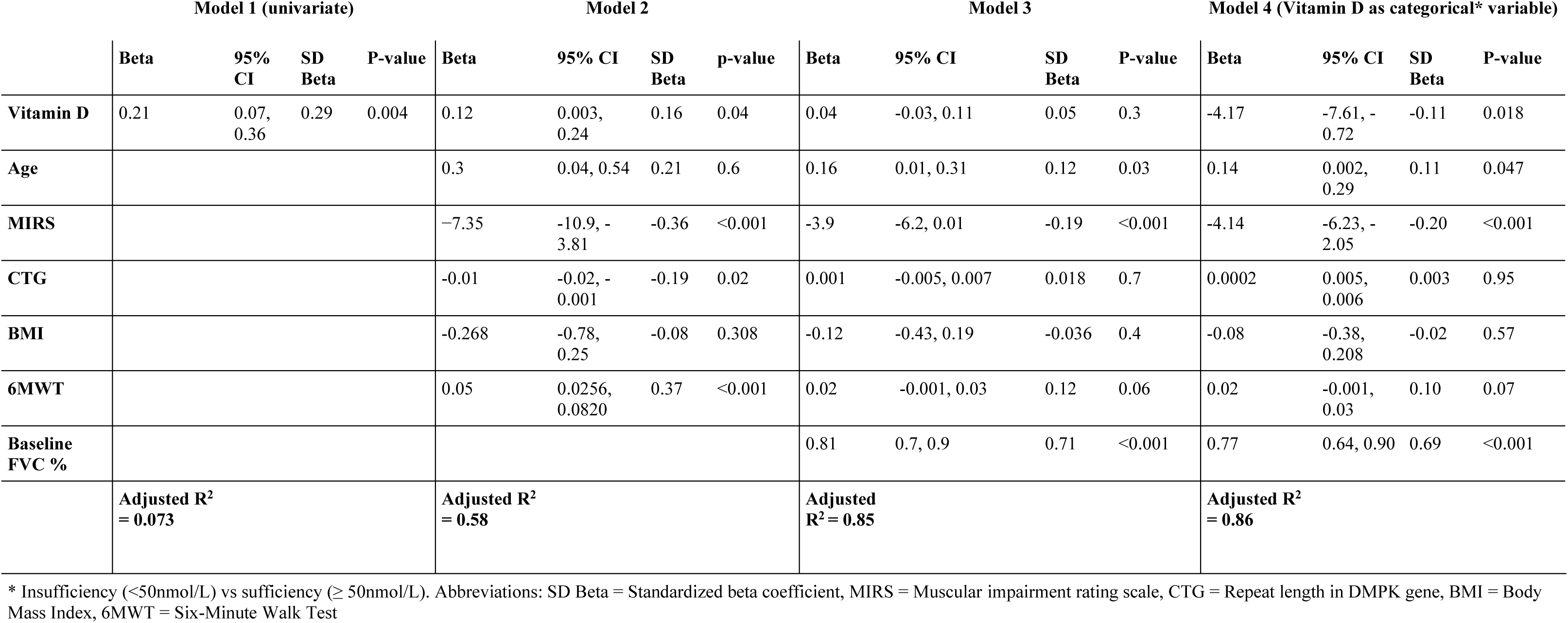
Regression models showing effect of baseline Vitamin D on follow-up FVC %.

|  | **Model 1 (univariate)** |  |  |  | **Model 2** |  |  |  | **Model 3** |  |  |  | **Model 4 (Vitamin D as categorical\* variable)** |  |  |  |
|---|---|---|---|---|---|---|---|---|---|---|---|---|---|---|---|---|
|  | **Beta** | **95% CI** | **SD Beta** | **P-value** | **Beta** | **95% CI** | **SD Beta** | **p-value** | **Beta** | **95% CI** | **SD Beta** | **P-value** | **Beta** | **95% CI** | **SD Beta** | **P-value** |
| **Vitamin D** | 0.21 | 0.07, 0.36 | 0.29 | 0.004 | 0.12 | 0.003, 0.24 | 0.16 | 0.04 | 0.04 | -0.03, 0.11 | 0.05 | 0.3 | -4.17 | -7.61, -0.72 | -0.11 | 0.018 |
| **Age** |  |  |  |  | 0.3 | 0.04, 0.54 | 0.21 | 0.6 | 0.16 | 0.01, 0.31 | 0.12 | 0.03 | 0.14 | 0.002, 0.29 | 0.11 | 0.047 |
| **MIRS** |  |  |  |  | −7.35 | -10.9, -3.81 | -0.36 | <0.001 | -3.9 | -6.2, 0.01 | -0.19 | <0.001 | -4.14 | -6.23, -2.05 | -0.20 | <0.001 |
| **CTG** |  |  |  |  | -0.01 | -0.02, -0.001 | -0.19 | 0.02 | 0.001 | -0.005, 0.007 | 0.018 | 0.7 | 0.0002 | 0.005, 0.006 | 0.003 | 0.95 |
| **BMI** |  |  |  |  | -0.268 | -0.78, 0.25 | -0.08 | 0.308 | -0.12 | -0.43, 0.19 | -0.036 | 0.4 | -0.08 | -0.38, 0.208 | -0.02 | 0.57 |
| **6MWT** |  |  |  |  | 0.05 | 0.0256, 0.0820 | 0.37 | <0.001 | 0.02 | -0.001, 0.03 | 0.12 | 0.06 | 0.02 | -0.001, 0.03 | 0.10 | 0.07 |
| **Baseline FVC %** |  |  |  |  |  |  |  |  | 0.81 | 0.7, 0.9 | 0.71 | <0.001 | 0.77 | 0.64, 0.90 | 0.69 | <0.001 |
|  | **Adjusted R<sup>2</sup> = 0.073** |  |  |  | **Adjusted R<sup>2</sup> = 0.58** |  |  |  | **Adjusted R<sup>2</sup> = 0.85** |  |  |  | **Adjusted R<sup>2</sup> = 0.86** |  |  |  |

\* Insufficiency (<50nmol/L) vs sufficiency (≥ 50nmol/L). Abbreviations: SD Beta = Standardized beta coefficient, MIRS = Muscular impairment rating scale, CTG = Repeat length in DMPK gene, BMI = Body Mass Index, 6MWT = Six-Minute Walk Test

### Multivariable models with Vitamin D as categorical predictor

Additionally, we examined the effect of vitamin D on FVC % as a categorical predictor comparing those with sufficient levels (≥50 nmol/L) vs those with insufficiency (<50 nmol/L). In this model, adjusting for age, MIRS, CTG expansion size, BMI, 6MWT and baseline FVC%, we found that vitamin D insufficiency was significantly associated with FVC% at follow-up: β = -4.17 (p > 0.018), indicating that patients with vitamin D insufficiency had on average 4.17 percentage-points lower FVC% at follow-up compared to those with sufficient levels.

## Discussion

In this prospective cohort study of adults with DM1, we found a significant association between baseline vitamin D status and FVC% at follow-up. The unadjusted association was relatively strong, indicated for example by the fact that patients with insufficient vitamin D levels (<50 nmol/L) had a 22 % higher absolute risk of worsening FVC% at follow-up. The association between baseline vitamin D and follow-up FVC% remained statistically significant after adjusting for multiple confounders. When including baseline FVC% in the model, i.e assessing change in FVC%, vitamin D remained statistically significant when tested as a categorical variable (i.e sufficiency vs insufficiency) but not when tested as continuous variable.

Although respiratory impairment is a major cause of morbidity and mortality in DM1 few longitudinal studies have examined risk factors that are potentially modifiable, and the relationship between Vitamin D and respiratory function has, to our knowledge, so far not been studied in this patient population^3, 19–21^. This knowledge gap is of clinical importance because effective strategies to alter long-term pulmonary trajectories in DM1 are currently limited.

To better understand the clinical relevance of our findings, it is important to consider the strength of the association between Vitamin D status and FVC% as well as the extent to which the association was independent of established determinants of respiratory function in DM1. Although the crude association between vitamin D status and respiratory function was relatively strong, part of this observed association could have been explained by the confounding effects of disease severity; as indicated by the fact that the effect estimate for vitamin D decreased after adjusting for MIRS, CTG-repeat length, age and BMI. However, despite this, vitamin D remained statistically significant and thus independently associated with FVC%. Analyzing vitamin D as a categorical variable, in the fully adjusted model, showed that patients with vitamin D insufficiency had on average 4.2 percentage-points lower FVC% at follow-up compared to those with sufficient levels. In other studies of lung disease the minimal clinically important difference (MCID) for FVC% has been estimated to 2-6%.^22, 23^ When interpreting these results it is important to consider the progressive nature of DM1 and the fact that even seemingly small short-term effect sizes can potentially accumulate over time and contribute to meaningful long-term differences in respiratory status. Furthermore, our regression models included MIRS and 6MWT which might lie on the causal pathway between vitamin D and respiratory function. This entails a risk of over-adjustment i.e attenuation of the true effect of vitamin D on FVC%. But perhaps more importantly, vitamin D insufficiency is easily detected and readily correctable with safe and inexpensive supplementation, without major risks of adverse effects. Because of the importance of preventing respiratory decline in DM1 the threshold for clinical action regarding vitamin D supplementation is low. Therefore, we believe that even estimates that appear to be conservative warrant clinical attention.

Nevertheless, it is worthwhile to emphasize that, regardless of vitamin D status, skeletal muscle weakness is an important factor influencing respiratory function in DM1, and both respiratory and peripheral muscle strength may contribute to pulmonary function and overall functional capacity. Therefore, individualized multidisciplinary rehabilitation programs incorporating aerobic and resistance exercise, as well as respiratory muscle training, and targeting other risk factors such as obesity and lifestyle factor, remain the cornerstone of management of respiratory impairment in this population.

When interpreting the results of this study, several methodological considerations are important to note. When vitamin D was examined as a categorical variable (deficiency vs. sufficiency), it remained statistically significant in the fully adjusted model, whereas the association was attenuated and no longer statistically significant when vitamin D was modelled as a continuous variable. A plausible explanation for this discrepancy is a threshold effect, where the influence of vitamin D on respiratory muscle function becomes most pronounced at deficient levels (<50 nmol/L), rather than following a linear dose-response relationship across the entire range of values.

Another important consideration is that vitamin D status was associated with FVC% at follow-up independently of disease severity, but is association with progression (i.e change in respiratory function) remains uncertain. FVC% changed relatively little during the study (From 65.7% to 63.6%) meaning that pulmonary function at follow-up was mostly determined by baseline status, making it difficult to detect independent predictors of progression. In this study, we chose to analyze change in FVC% using a baseline adjusted linear model because of its advantages over change-score analysis^24^, but it is possible that other methods such as structural equation modeling (SEM) would have yielded more accurate results. Nevertheless, because of the modest change in FVC%, this study lacked power to produce precise estimates on the long-term consequences of vitamin D insufficiency, possibly regardless of statistical method. These findings are themselves important though for future studies as they highlight the need for a longer follow-up than 4 years. Additional analyses characterizing the overall change in respiratory function and its relationship with demographic characteristics have been made and will be presented separately.

Additional limitations of this study were the single-centre design, and the fact that it was conducted in an area with more limited sun light than many other countries, which may limit generalizability. We did not account seasonal variation in the analyses and were unable to control for vitamin D supplementation. Although a high level of vitamin D supplementation would likely have weakened any associations. Also, as shown in our causal diagram, seasonal variation influences vitamin D levels, but does not affect long-term lung function and can therefore not have a confounding effect^25^. However, it could still be of relevance because it could interact with other factors such as reduced mobility, the effect of which could be more pronounced in geographical areas with more sunlight. Another limitation is that we did not have data on use of non-invasive ventilation which is one of the most important aspects regarding respiratory health in DM1. Sex differences in vitamin D metabolism, including the higher prevalence of vitamin D deficiency observed among men in our cohort, may represent an additional source of variation; however, sex was not independently associated with follow-up FVC% in our analysis. Residual confounding by unmeasured or imperfectly measured indicators of disease severity cannot be excluded, particularly given the multisystem nature of DM1. Also, the follow-up period, while clinically meaningful, did not fully capture longer-term trajectories of respiratory decline in this slowly progressive disease. Finally, perhaps the most important limitation of the present study is that it cannot address the clinically most relevant question: whether or not correction of vitamin D insufficiency can alter the trajectory of respiratory decline in patients with DM1. While our findings demonstrate a significant association between lower vitamin D levels and impaired respiratory function, our findings alone cannot determine the causal effects of Vitamin D supplementation.

The main strengths of this study include the prospective and longitudinal design which ensures clear temporal ordering between exposure and outcome. Second, the sample size is relatively large for a rare neuromuscular disorder such as DM1, enhancing the precision of the estimates and increasing the robustness of the findings. Third, the longitudinal assessment of FVC enables evaluation of within-individual change over time, addressing a gap in the literature where prospective cohort studies in DM1 remain scarce. Fourth, all examinations were conducted at a specialized neuromuscular center by highly trained personnel using standardized protocols, reducing measurement variability and strengthening internal validity. Fifth, the systematic and uniform data collection across participants limits information bias and supports the reliability of the observed associations.

In conclusion, the findings in this prospective cohort study suggests that vitamin D insufficiency is associated with lower FVC% in adults with DM1, with potential implications for long-term respiratory trajectories. These findings highlight the need to further study the relationship between vitamin D status and lung function over time and examine whether optimizing vitamin D status could improve respiratory health in this population.

## Data Availability

Data produced in the present study are not available upon request due to legal and regulatory restrictions imposed by Swedish law and the European Union

## Supplementary material

Figure caption: Supplementary figure. Hypothesized causal relationships illustrated in a directed acyclic graph (DAG)

## Declaration of generative AI

During the preparation of this work the first author used generative AI [ChatGPT] for language editing and code review [Claude]. All authors reviewed and edited the content and take full responsibility for the content of the published article

## Author contributions

Daniel Jaraj participated in the data collection, statistical analyses, interpretation of the data and drafting and revising the manuscript. Ólöf Elíasdóttir participated in the data collection, interpretation of the data and revising the manuscript. Stefan Winblad participated in the study design, interpretation of the data and revising the manuscript. Christopher Lindberg conceived the study, participated in the design and collection of data, interpretation of the data and revising the manuscript

## Statements and declarations

### Ethical considerations

The study was conducted in accordance with the Declaration of Helsinki (2000) and was approved by The Swedish Ethical Review Authority (number 826-16).

### Consent to participate

Informed consent (written and verbal) was collected from all participants prior to entering the study.

### Consent for publication

Not applicable

